# Severity-stratified, longitudinal cis-eQTL mapping enables high-sensitivity detection of candidate causal genes for critical COVID-19

**DOI:** 10.64898/2026.10.14.26365529

**Authors:** Ernie Chen, Gisela Gabernet, David B. Corry, Farrah Kheradmand, Lindsey R. Baden, Rafick-Pierre Sekaly, Grace A. McComsey, Elias K. Haddad, Charles B. Cairns, Nadine Rouphael, Ana Fernandez-Sesma, Viviana Simon, Jordan P. Metcalf, Nelson I. Agudelo Higuita, Catherine L. Hough, William B. Messer, Mark M. Davis, Kari C. Nadeau, Bali Pulendran, Holden T. Maecker, Monica Kraft, Chris Bime, Elaine F. Reed, Joanna Schaenman, David J. Erle, Carolyn S. Calfee, Mark A. Atkinson, Scott C. Brakenridge, Esther Melamed, Lauren I. R. Ehrlich, Albert C. Shaw, David A. Hafler, Alison D. Augustine, Patrice M. Becker, Bjoern Peters, Al Ozonoff, Joann Diray-Arce, Ofer Levy, Steven E. Bosinger, Walter Eckalbar, IMPACC Network, Ruth R. Montgomery, Ira M. Hall, Steven H. Kleinstein, Dylan Duchen

## Abstract

Host genetic variation influences heterogeneity in infectious disease severity. Genome-wide association studies (GWAS) have identified 69 lead single nucleotide polymorphisms (SNPs) associated with critical COVID-19, but most lack known regulatory mechanisms. To date, the genetic cis-regulatory effects of critical COVID-19 GWAS lead SNPs have not been systematically characterized across acute illness and convalescent phases in a COVID-19 specific cohort. Here, performing severity-stratified, longitudinal cis-expression quantitative trait loci (cis-eQTL) mapping and colocalization with critical COVID-19 GWAS signals, we find that host cis-regulatory effects vary across clinical severity and disease phase. We analyzed peripheral blood mononuclear cells from 1,031 patients in the Immunophenotyping Assessment in a COVID-19 Cohort (IMPACC), a prospective multicenter study of adults hospitalized with COVID-19, profiled across 10 timepoints spanning acute illness and convalescence. We identified 23 GWAS lead SNPs that colocalized with cis-eQTLs at 26 candidate causal genes, with 16 detectable only when severity stratification and longitudinal sampling were incorporated. A colocalized, protective allele (chr19:4,717,660) at the *TMIGD2* locus upregulated *TMIGD2* expression exclusively during the acute phase in the prolonged-critical/fatal cohort, with no detectable effect in the rapid-recovery cohort. Our results show that host genetic effects on critical COVID-19 can be context-dependent, with cis-regulatory effects varying across clinical severity and disease phase. Severity-stratified, longitudinal cis-eQTL designs have greater sensitivity to identify candidate causal genes for critical COVID-19 GWAS associations than single-timepoint cross-sectional studies.

**Significance Statement:** Genome-wide association studies have identified 69 loci associated with life-threatening COVID-19, but most lead SNPs, the strongest association signal at each locus, lie outside protein-coding regions and act via unknown effects on gene regulation. We analyzed paired genetic and gene expression data from 1,031 hospitalized COVID-19 patients in IMPACC, sampled across ten timepoints and stratified by clinical severity. This analysis linked 23 of these risk variants to candidate genes, more than doubling the single-timepoint yield. Many effects appeared only in specific severity cohorts or timepoints. In critically ill patients early in hospitalization, the GWAS-associated allele increased expression of *TMIGD2*, a costimulatory receptor that amplifies T cell activation and may support antiviral responses. These variant-gene pairs warrant functional validation.

## Introduction

Host genetic variation is a major determinant of clinical heterogeneity in infectious disease (1, 2). This is particularly evident in Coronavirus Disease 2019 (COVID-19), caused by the SARS-CoV-2 virus. While the majority of COVID-19 cases are mild, ∼5-6% of infections progress to critical illness prompting hospitalization and intensive care, with significant variability in clinical outcomes even among hospitalized patients (3, 4). Critical COVID-19 cases follow a distinct pathophysiology compared to mild or moderate illness, such as dysregulated type I interferon responses, excessive inflammation, and vascular dysfunction (5, 6). Seminal studies early in the pandemic demonstrated that this clinical heterogeneity is partially driven by host genetics (7). While rare, deleterious loss-of-function (LOF) variants explain ∼3.5% of life-threatening COVID-19 cases (8), the majority remain genetically complex with no clear familial inheritance pattern (9). Large-scale meta-analyses of critical COVID-19 genome-wide association studies (GWAS) have identified 69 lead genetic variants associated with critical illness (10–12). However, the biological mechanisms by which these variants influence disease severity remain incompletely understood.

Most GWAS variants lie in non-coding regions of the genome and are presumed to influence disease risk by altering gene expression (13). An established approach for linking GWAS variants to their target genes is cis-expression quantitative trait locus (cis-eQTL) mapping, which tests whether a risk variant is associated with expression of a nearby gene (14, 15). Existing cis-eQTL reference datasets, however, are poorly positioned to study the biology of critical COVID-19. Most derive from healthy donors or cohorts with mixed disease states (14), so their eQTL estimates reflect a population average rather than gene regulation specific to a disease context. Moreover, many regulatory effects are observed only by immune activation: over half of cis-eQTLs in monocytes and macrophages are detected only after stimulation (16, 17). A population average cis-eQTL map therefore obscures the genetic regulation most relevant to critical COVID-19 and limits causal inference about the target genes of GWAS variants (18). Beyond this context-specificity gap, existing reference datasets also lack sampling across the disease course of critical COVID-19. Because gene expression is dynamic throughout illness while genotype is not, these datasets cannot reveal how host genetic regulation varies with disease progression (19). Consequently, existing cis-eQTL maps offer only a static view of how variants influence gene expression during COVID-19. A cis-eQTL analysis conducted in a critical COVID-19 patient cohort is therefore essential to identify the target genes through which these variants shape disease severity. A longitudinal design would further reveal how GWAS variant effects on gene expression evolve from acute illness to convalescence.

To characterize how host genetic regulation changes over the course of critical illness, we leveraged the Immunophenotyping Assessment in a COVID-19 Cohort (IMPACC), a longitudinal study of 1,164 hospitalized adults with laboratory-confirmed SARS-CoV-2 infection (20). Prior IMPACC reports defined clinical trajectory groups (TGs) and multi-omic correlates of acute severity and recovery (20–23), providing validated severity phenotypes for cis-eQTL analyses. Genetic cis-regulatory effects act upstream of clinical outcome, so studying critical COVID-19 GWAS variants throughout the course of hospitalization can reveal how genetic regulation contributes to the divergence between rapid recovery and progression to critical illness. We first performed unstratified cis-eQTL analysis from paired peripheral-blood mononuclear-cell (PBMC) RNA-seq data and matched genotypes at hospital admission. We then expanded this analysis across two dimensions by mapping cis-eQTLs stratified by clinical severity cohorts and across ten timepoints spanning acute illness and convalescence. This severity-stratified longitudinal design detected many critical COVID-19 risk variants that exhibit regulatory effects specific to disease phases and clinical severity TGs, providing fresh insight into cis-eQTL associations with COVID-19 severity and outcome.

## Results

### Severity-stratified, longitudinal analysis reveals context-dependent genetic regulation of critical COVID-19

The cis-regulatory effects of critical COVID-19 GWAS lead SNPs during acute illness have not been systematically characterized in a COVID-19 exclusive cohort. We first addressed this by asking which of the 69 lead SNPs identified by meta-analyses (11, 12) function as cis-eQTLs at hospital admission with COVID-19. We leveraged IMPACC, a longitudinal cohort of 1,164 hospitalized COVID-19 patients (21). We performed linear cis-eQTL mapping at hospital admission (Visit 1; within 48 hours of admission) using data from 921 IMPACC participants who passed genotype QC and had matched PBMC RNA-seq data (out of 1,031 passing genotype QC). We evaluated 58 QC-passing lead SNPs (of 69) against the 887 protein-coding genes lying within ±1 Mb of any lead SNP, adjusting for age, sex, and principal components derived separately from expression and genotype data (see Methods). Significant cis-eQTLs were defined as lead SNP-gene pairs with eigenMT-adjusted P < 0.01, correcting per-gene for the effective number of independent SNPs tested within the ±1 Mb cis-window (24). This screen identified 61 significant SNP-gene pairs involving 32 of 58 lead SNPs (55%) and 52 of 887 genes (5.9%) (Fig. 2). The remaining 45% of lead SNPs showed no detectable cis-regulatory effect at hospital admission.

**Fig. 1.**
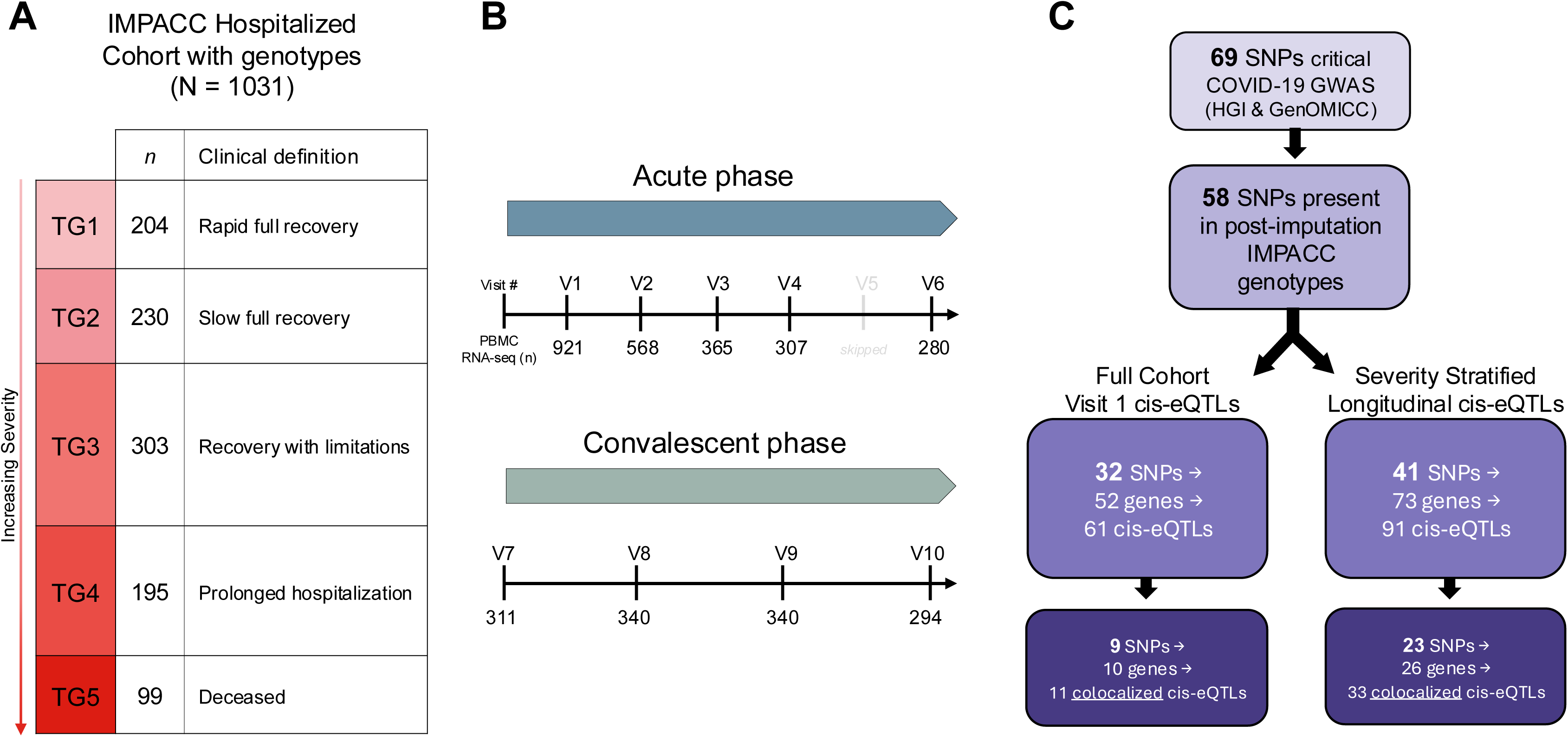
Severity-stratified, longitudinal cis-eQTL design implicates context-dependent regulatory effects in critical COVID-19. (A) Severity stratification of the genotyped IMPACC cohort (n = 1,031). Participants were classified into five trajectory groups (TG) based on longitudinal WHO severity scores, ranging from rapid recovery (TG1, n = 204) to death (TG5, n = 99), with increasing severity indicated by color intensity from light pink to dark red. For cis-eQTL analyses, groups were aggregated into rapid-recovery (TG1-3) and prolonged-critical/fatal (TG4-5) cohorts. (B) Timeline of biospecimen sampling spanning acute (V1-V6, Days 1-28 post-admission; V5 excluded due to low sample size) and convalescent (V7-V10, Months 3-12 post-discharge) phases. Numbers below the timeline indicate the count of PBMC RNA-seq samples available at each timepoint; attrition reflects hospital discharge or mortality. (C) Schematic of the analytical workflow. Starting from 69 established critical COVID-19 GWAS lead SNPs (HGI C GenoMICC) (11, 12), 58 were available for cis-eQTL analysis in IMPACC. Parallel workflows comparing unstratified, single-timepoint analysis at hospital admission (V1, left) versus severity-stratified, longitudinal analysis (right). Darkest purple boxes indicate the count of variants showing Bayesian colocalization (PP.H4 > 0.5) with the GenOMICC critical illness GWAS signals (11).

**Fig. 2.**
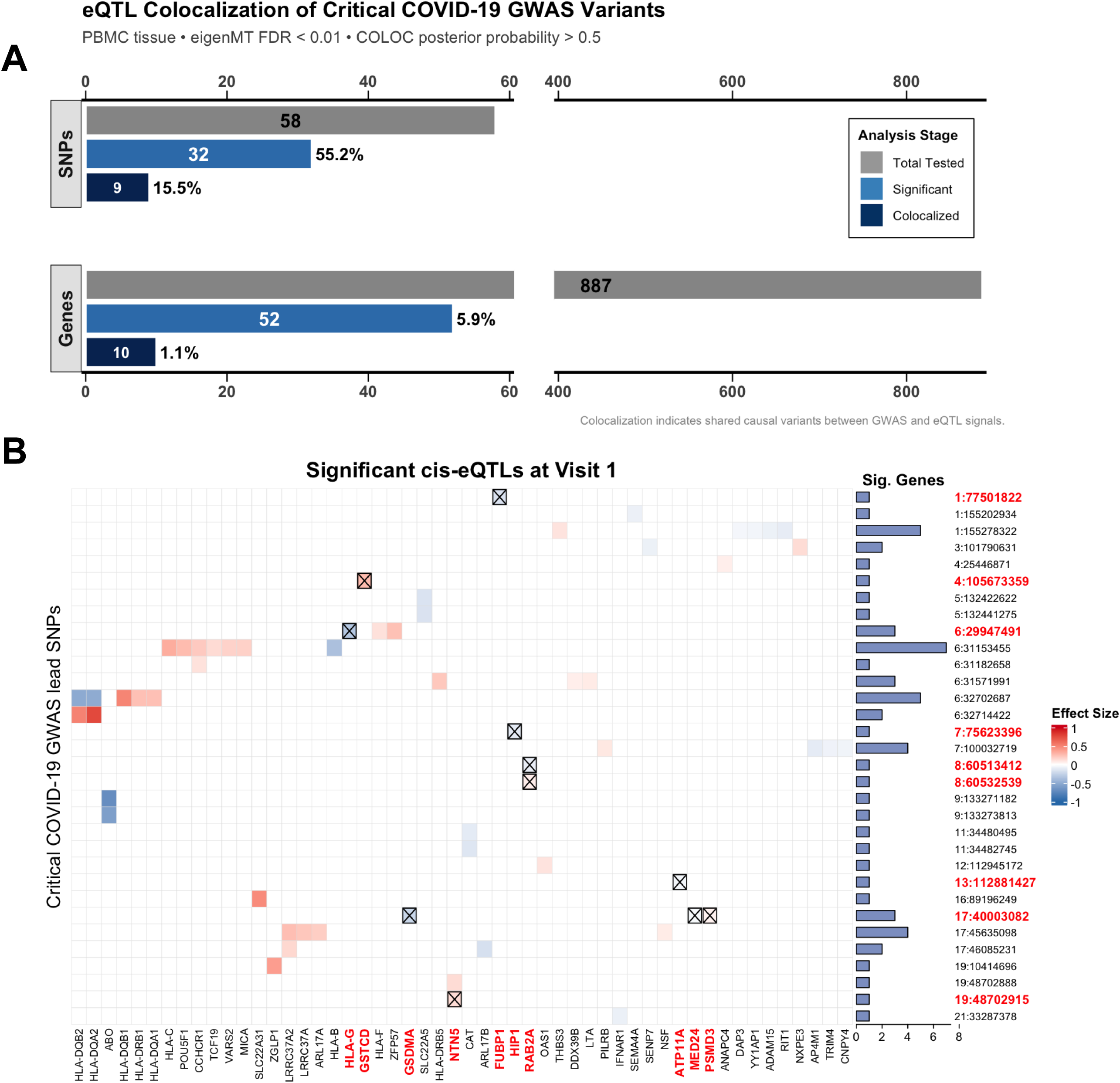
cis-eQTL mapping at hospital admission identifies regulatory effects for critical COVID-19 GWAS lead SNPs. (A) Attrition of GWAS lead SNPs and candidate causal genes at hospital admission. Top set of bars shows all reported GWAS lead SNPs (gray, n = 58), those with at least one significant cis-eQTL activity (light blue, n = 32), and those colocalizing with critical COVID-19 GWAS signals (dark blue, n = 9). Lower bars track the corresponding attrition of protein-coding genes within ±1 Mb windows of lead SNPs (gray, n = 887; significant cis-eQTL associations, n = 52; candidate causal genes from colocalization, n = 10). (B) Heatmap of significant SNP-gene cis-eQTL associations at hospital admission (n = 921). Rows represent GWAS lead SNPs with at least one significant cis-eQTL association, ordered by chromosome:position (GRCh38); columns represent the associated genes. Cell color indicates the effect size (β) of the effect allele (red = upregulation; blue = downregulation). SNP-gene pairs showing Bayesian colocalization (PP.H4 > 0.5) are marked with crossed cells (X) and have bold red axis labels. The horizontal bar chart (right) shows the number of genes with significant cis-eQTL associations at each lead SNP locus.

The absence of detectable cis-regulatory activity for a substantial fraction of lead SNPs raised the possibility that their effects emerge at specific clinical severity trajectories and/or disease phases. To test this, we extended our cis-eQTL analysis across IMPACC’s clinical trajectory groups (TGs) and longitudinal timepoints. IMPACC participants were previously assigned to one of five clinical TGs (TG1 through TG5) by unsupervised clustering of longitudinal respiratory ordinal scores over the first 28 days of hospitalization, ranging from TG1 (brief hospital stay) to TG5 (fatal outcome within 28 days) (21). For all downstream analyses, we combined TG1-3 into a rapid recovery cohort (n = 737 across the full genotyped cohort) and TG4-5 into a prolonged critical/fatal cohort (n = 294). PBMC RNA-seq data were collected at 10 scheduled timepoints spanning acute hospitalization through convalescence, including six acute visits (Visits 1 through 6, hereafter V1-V6; corresponding to days 1, 4, 7, 14, 21, 28 post-admission) and four subsequent convalescent visits (V7-V10; months 3, 6, 9, and 12 post-discharge) (20, 21). We analyzed nine of these timepoints (V1-V4, V6-V10), excluding V5 where only 98 samples had matched genotype and PBMC RNA-seq data, of which only 10 were from TG1-3 patients, insufficient for reliable cis-eQTL detection. TG4-5 samples at V5 were excluded in parallel to maintain matched trajectory comparisons across visits. Per-visit sample sizes varied based on RNA-seq availability, ranging from 660 (TG1-3 at V1) to 41 (TG4-5 at V10).

This design yielded 18 independent cis-eQTL analyses (2 severity groups × 9 visits), adding clinical severity and disease phase as analytical dimensions beyond our hospital admission analysis. This expanded analysis raised the fraction of lead SNPs with detectable cis-regulatory activity from 32 of 58 SNPs (55%) at hospital admission to 41 (71%) across all 18 strata (Fig. 2). In total, we detected 91 significant SNP-gene pairs involving 73 of 887 genes (8.2%). Eleven lead SNPs showed cis-regulatory activity that were not detected in our hospital admission-only analysis, while two lead SNPs detected as cis-eQTLs in our hospital admission analysis (chr4:105,673,359 and chr13:112,881,427) showed no significant cis-regulatory activity in any of the 18 severity-by-visit analyses. Rapid recovery patients (TG1-3) had more than twice as many cis-eQTLs per visit compared to the critical/fatal subset of patients (TG4-5). Together, the 11 additional lead SNPs detected across the expanded analysis indicated that regulatory activity at critical COVID-19 loci varies with both clinical severity and disease phase.

### Colocalization across severity-stratified, longitudinal cis-eQTLs identifies phase- and severity-specific regulation at critical COVID-19 loci

To determine whether the cis-eQTLs detected in our hospital admission analysis share causal variants with the underlying critical COVID-19 GWAS associations, we performed Bayesian colocalization analysis (25). Colocalization estimates the posterior probability (PP.H4) that two association signals share a causal variant, nominating the corresponding gene as a candidate causal gene for critical COVID-19. We first applied this framework to our 61 hospital admission cis-eQTL pairs, using summary statistics from the GenOMICC critical COVID-19 meta-GWAS (n = 23,707 critical illness cases excluding 23andMe) (11), whose case definition (requiring respiratory support or COVID-19-related death) matched the critical illness phenotype of all IMPACC participants at enrollment (10, 11). At a posterior probability threshold of 0.5, consistent with the threshold applied in the GenOMICC GWAS (11), 9 of 58 GWAS lead SNPs colocalized with cis-eQTLs involving 10 genes. The remaining 49 SNPs (84%) fell into two categories: 26 lacked detectable cis-regulatory activity at hospital admission, and 23 had a significant cis-eQTL but did not colocalize with the GWAS signal. Colocalization of hospital admission cis-eQTLs alone therefore nominated candidate causal genes for only 16% of critical COVID-19 lead SNPs.

We next applied this colocalization framework to the 91 cis-eQTL pairs (eigenMT-adjusted P < 0.01) detected across all 18 severity-by-visit analyses. Of the 41 lead SNPs with significant cis-regulatory effects in this expanded analysis, 23 colocalized with cis-eQTLs at 26 genes (33 SNP-gene pairs total). Of these 23 lead SNPs, 7 had also colocalized in the hospital admission analysis, while 16 were newly detected through the severity-stratified, longitudinal analysis. Two hospital admission colocalizations did not appear in the expanded analysis. Among the 33 colocalized SNP-gene pairs, 6 were detected only in TG4-5, 16 only in TG1-3, and 11 in both cohorts (Fig. 3B). Two thirds of colocalizations were detected in only one severity cohort, suggesting that genetic regulation at these loci varies with clinical context. Across all 58 critical COVID-19 lead SNPs tested, our hospital admission analysis identified candidate causal genes for 9 (16%), whereas the severity-stratified, longitudinal analysis identified candidates for 23 (40%), including 16 loci with no detectable colocalization in the former analysis. This difference in detection was statistically significant (McNemar exact test, P = 0.0013). The remaining 35 SNPs (60%) showed no PBMC cis-eQTL colocalization: 17 lacked any significant cis-eQTL across the severity-stratified, longitudinal analysis, and 18 had significant cis-eQTLs that did not colocalize with their corresponding GWAS signal.

**Fig. 3.**
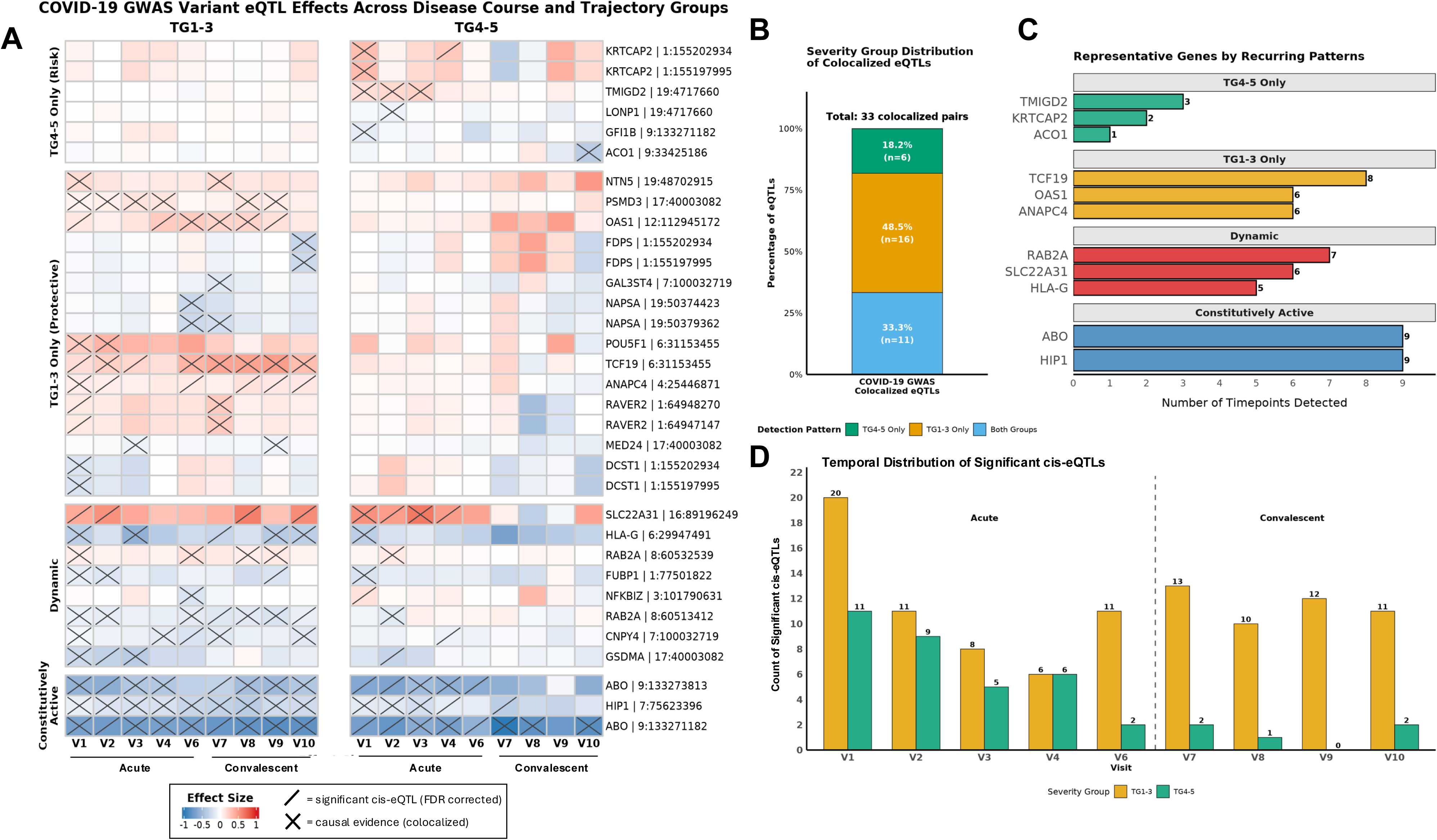
Severity-stratified, longitudinal cis-eQTL analysis identifies context-dependent regulatory effects in critical COVID-19. (A) Heatmaps of cis-eQTL effects across disease course (V1-10, excluding V5) stratified by severity cohort. The left panel shows effects in the rapid-recovery cohort (TG1-3); the right panel shows effects in the prolonged-critical/fatal cohort (TG4-5). Rows represent SNP-gene pairs grouped by similar regulatory patterns (e.g., “TG4-5 Only”). Cell color indicates effect size (β) of the effect allele (red = upregulation; blue = downregulation). A diagonal slash (/) indicates a statistically significant cis-eQTL association (eigenMT-adjusted FDR < 0.01). Crossed cells (X) indicate Bayesian colocalization (PP.H4 > 0.5) at that specific severity-by-visit stratum. (B) Distribution of the 33 colocalized SNP-gene pairs by severity cohort detection (TG4-5 only, TG1-3 only, or both). (C) Representative genes illustrating each regulatory pattern. Bars represent the number of timepoints at which a significant cis-eQTL association was detected for each gene. Green bars indicate severity-cohort-specific regulation in TG4-5; gold bars indicate severity-cohort-specific regulation in TG1-3; red bars indicate dynamic regulation in both cohorts with distinct temporal profiles; blue bars indicate constitutive regulation across both cohorts and all timepoints. (D) Temporal distribution of significant cis-eQTLs stratified by severity cohort. Bars represent the count of SNP-gene pairs with significant cis-eQTL association (eigenMT-adjusted FDR < 0.01) at each visit, stratified by severity cohort (gold = TG1-3; green = TG4-5). The dashed line separates the acute phase (V1-V6, V5 excluded) from the convalescent phase (V7-V10).

### Representative loci exhibit distinct severity- and time-dependent regulatory patterns

Having identified 33 colocalized SNP-gene pairs across the severity-stratified, longitudinal analysis, we next asked whether their regulatory effects had observable recurring patterns across severity cohorts and disease phases. We identified four such patterns: signals specific to TG4-5 (e.g., *TMIGD2*, *LONP1, KRTCAP2*); signals specific to TG1-3 (e.g., *OAS1*, *PSMD3*, *TCF19*); dynamic signals present in both cohorts but with distinct temporal profiles (e.g., *RAB2A*, *SLC22A31*, *HLA-G*); and constitutive signals detected across visits and cohorts (e.g., *ABO*, *HIP1*) (Fig. 3A). Of the 11 SNP-gene pairs detected in both TG4-5 and TG1-3, 8 (dynamic) showed distinct temporal profiles and 3 (constitutive) were detected across all visits (Fig. 3B).

To illustrate how the four regulatory patterns identified above manifest at individual loci, we examined one representative gene per pattern: *TMIGD2* (TG4-5-specific), *OAS1* (TG1-3-specific), *HLA-G* (asymmetrically dynamic), and *ABO* (constitutive). At each locus, the protective and risk alleles are the alleles associated with lower and higher odds of critical COVID-19, respectively, in the GenOMICC GWAS. At the lead SNP nearest *TMIGD2* (chr19:4,717,660; protective allele A), encoding the costimulatory receptor CD28H (26), the protective allele was associated with upregulation of *TMIGD2* detectable exclusively in TG4-5 during the early acute phase (V1-3; peak β = +0.22, eigenMT-adjusted P < 0.01), with no detectable effect in TG1-3 or at later timepoints in TG4-5 (Fig. 4A, top). The same lead SNP also colocalized with downregulation of the adjacent *LONP1* (mitochondrial Lon protease) (27) in TG4-5 at V2 (β = −0.08, eigenMT-adjusted P < 0.001), implicating the same locus in opposite-direction regulation of two neighboring genes during early acute illness.

**Fig. 4.**
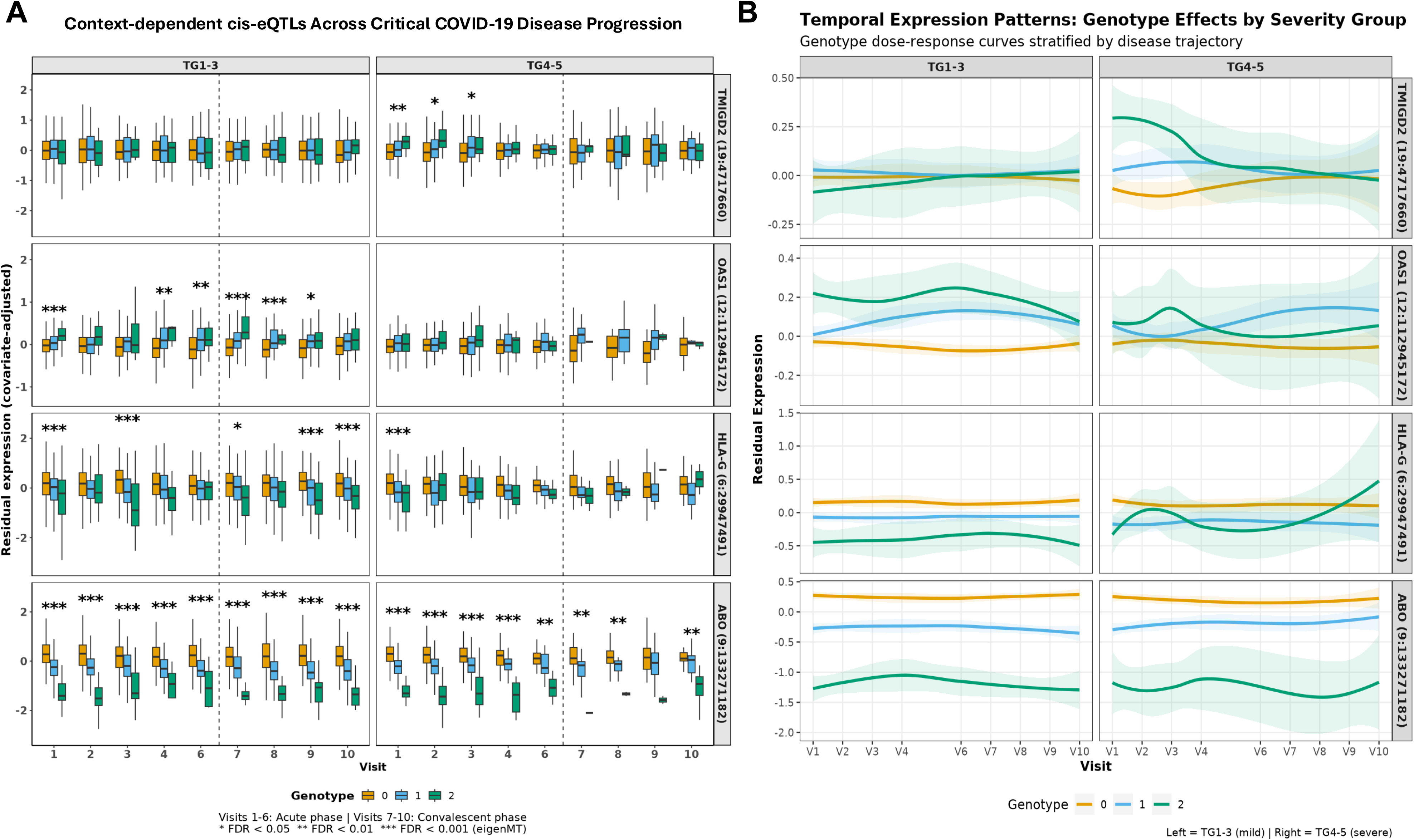
Severity-stratified, longitudinal expression of representative cis-regulatory loci. (A) Temporal dynamics of cis-regulatory effects for selected loci illustrating the regulatory patterns observed across colocalizations. Boxplots show covariate-adjusted residual gene expression at each visit, stratified by severity trajectory (left: TG1-3, rapid-recovery; right: TG4-5, prolonged-critical/fatal) and genotype dosage (0 = reference allele homozygote, gold; 1 = heterozygote, blue; 2 = effect allele homozygote, green). Asterisks denote the significance of the cis-eQTL association at each specific visit-trajectory stratum (*eigenMT-adjusted FDR < 0.05; **<0.01; ***<0.001). Dashed vertical lines separate acute (Visits 1-4, 6) from convalescent (Visits 7-10) phases. (B) Loess-smoothed temporal dose-response curves illustrating genotype effects across severity trajectories. Shaded regions represent 95% confidence intervals. *TMIGD2* (top) displays protective-allele-dependent upregulation restricted to the acute phase (V1-V3) of TG4-5; *OAS1* displays protective-allele-dependent upregulation restricted to TG1-3 across both acute and convalescent phases. *HLA-G* shows the dynamic regulatory pattern, with cis-regulatory effects detectable in both cohorts but with distinct temporal profiles. ABO (bottom) exhibits constitutive regulation, with distinct genotype effects persisting across all timepoints and trajectories.

In contrast, *OAS1,* which encodes an antiviral effector at an established COVID-19 GWAS locus (28), illustrates the TG1-3-specific pattern. At the lead SNP nearest *OAS1* (chr12:112,945,172; protective allele C), the protective allele was associated with upregulation of *OAS1* detectable exclusively in TG1-3 across acute (V1, V4, and V6; peak β = +0.26, eigenMT-adjusted P < 0.001) and convalescent (V7-9; peak β = +0.22, eigenMT-adjusted P < 0.001) timepoints, with no detectable effect in TG4-5 (Fig. 4A, second row). The cis-regulatory effect is therefore preserved in patients with rapid recovery but undetectable in those with prolonged-critical/fatal illness.

*HLA-G,* which encodes an immune-modulatory MHC class I molecule (29), illustrates the asymmetrically dynamic pattern. At the lead SNP nearest *HLA-G* (chr6:29,947,491; risk allele C), the risk allele was associated with downregulation of *HLA-G* in TG1-3 across both acute (V1 and V3; peak β = −0.57, eigenMT-adjusted P < 0.001) and convalescent (V7, V9, and V10; peak β = −0.38, eigenMT-adjusted P < 0.001) phases. In TG4-5, the effect was limited to V1 (β = −0.36, eigenMT-adjusted P < 0.001) (Fig. 4A, third row). *ABO,* the most frequently replicated COVID-19 GWAS locus (30), illustrates the constitutive expression pattern. At the lead SNP nearest *ABO* (chr9:133,271,182; risk allele T), the risk allele was associated with consistent downregulation of *ABO* at 17 of 18 severity-by-visit strata (peak β = −0.76, eigenMT-adjusted P < 0.001) (Fig. 4A & 4B, bottom row). Together, these four loci indicate that critical COVID-19 GWAS SNPs exert cis-regulatory effects that vary across clinical severity and disease phase.

## Discussion

Here, we identify candidate causal genes for critical COVID-19 GWAS lead SNPs and show that host genetic regulation of these genes varies across clinical severity and disease phase. In a severity-stratified, longitudinal cis-eQTL analysis of 1,031 hospitalized COVID-19 patients, 23 GWAS lead SNPs colocalized with cis-eQTLs at 26 candidate causal genes (Fig. 3). Sixteen of these 23 colocalizations were detectable only when severity stratification and longitudinal sampling were incorporated, improving the yield more than two-fold relative to our unstratified hospital admission analysis.

The two additional dimensions of clinical severity and time significantly improved our colocalization rates from 16% (9/58) in an unstratified, single-timepoint analysis at hospital admission to 40% (23/58) in our severity-stratified, longitudinal analysis. The improvement addresses the colocalization gap, where most GWAS variants are not linked to candidate causal genes through cross-sectional cis-eQTLs alone (31). For context, the GTEx atlas reports a per-trait colocalization rate of 21% across single tissues (14). Our findings therefore demonstrate the empirical value of disease-relevant and longitudinal cis-eQTL designs for linking GWAS lead SNPs to candidate causal genes in critical COVID-19.

Among our candidate causal genes, *TMIGD2* illustrates both severity-cohort-specific and temporal regulation. The protective allele upregulated *TMIGD2* expression exclusively in severely ill TG4-5 patients during the early acute phase (V1-V3), with no detectable effect in the milder cases in TG1-3. TMIGD2 (CD28H) is a costimulatory receptor highly expressed on naive T cells and NK cells, where binding by its ligand HHLA2 amplifies cell activation, proliferation, and cytokine production through AKT signaling (26). We therefore hypothesize that the protective allele supports a stronger initial antiviral response by upregulating *TMIGD2* expression during the early acute window of critical illness. Prior IMPACC analyses reported higher frequencies of circulating T and NK cells at hospital admission in participants with rapid-recovery trajectories (TG1-3), and a decline in T cells and granzyme B-producing cytotoxic NK cells over the disease course in those with fatal outcome (TG5) (20), supporting the biological plausibility of this hypothesis. Costimulation-linked transcripts are also enriched in SARS-CoV-2-reactive CD8+ T cells of severely ill COVID-19 patients (32).

The ligand of TMIGD2, HHLA2, also binds an opposing coinhibitory receptor, KIR3DL3, which is upregulated on terminally differentiated CD8+ T cells and CD56^dim^ NK cells (33). KIR3DL3 binding by HHLA2 attenuates cytotoxic T cell and NK cell activation. Given this opposing receptor, *TMIGD2* upregulation by the protective allele may not only strengthen the initial costimulatory signal but also extend the productive activation window before KIR3DL3-mediated inhibition takes over, supporting a more sustained antiviral response. Testable therapeutic hypotheses arising from this finding include strengthening early TMIGD2-HHLA2 costimulation or blocking KIR3DL3-mediated inhibition during the early acute phase in critical patients carrying the risk allele. KIR3DL3-blocking antibodies developed for cancer immunotherapy selectively block KIR3DL3-mediated inhibition on activated CD8+ T cells and NK cells while preserving TMIGD2-HHLA2 costimulation (33). Nevertheless, the therapeutic implications of our *TMIGD2* finding remain hypothesis-generating, pending functional validation.

Whereas the *TMIGD2* cis-eQTL colocalization was detectable only in TG4-5, the *OAS1* colocalization was detectable only in TG1-3. The protective allele of *OAS1* upregulated expression in TG1-3 across both acute (V1, V4, V6) and convalescent (V7-V9) phases. *OAS1* encodes an interferon-induced enzyme that, upon detecting single-stranded viral RNA, activates ribonuclease L to degrade viral genomes during innate antiviral defense (28). The protective allele at this well established GWAS locus produces a longer, prenylated isoform with enhanced antiviral activity (34, 35). Our findings extend these reports by showing that the *OAS1* cis-regulatory effect is restricted to patients with rapid recovery (TG1-3) and is sustained from acute illness through convalescence.

The absence of a detectable cis-eQTL effect at any timepoint in TG4-5 is consistent with a context-dependent mechanism for *OAS1* protection. Interferon signaling is impaired in critical COVID-19 (8, 36). A cis-regulatory increase in *OAS1* expression, an interferon-induced gene, would have limited functional consequence if interferon signaling has already become dysregulated. The cis-regulatory effect on *OAS1* may therefore be detectable in patients with functional interferon responses but masked in patients whose interferon-pathway dysregulation alters the broader transcriptional context within which *OAS1* requires.

Beyond the severity-cohort-specific expression patterns illustrated by *TMIGD2* and *OAS1*, our colocalizations also showed dynamic and constitutive regulatory effects. *HLA-G*, a non-classical MHC class I molecule with immune-modulatory function (29), demonstrates a dynamic pattern; its cis-regulatory effect was sustained across multiple visits in TG1-3 but restricted to V1 in TG4-5. *ABO*, the most commonly replicated COVID-19 GWAS locus (30), illustrates the constitutive pattern, with cis-regulatory effects detectable across nearly all severity-by-visit strata. Together, these findings indicate that the cis-regulatory architecture of critical COVID-19 GWAS loci is non-uniform, ranging from constitutive activity to context-specific within particular severity cohorts and disease phases.

Our study has several limitations to consider. We anchored our colocalization analysis on 58 lead SNPs from published GWAS meta-analyses. Accordingly, association signals not identified by these GWAS were not tested for colocalization, so a regulatory effect driven by such a signal would not be detected by our approach. Bulk PBMC transcriptomics cannot resolve cell-type-specific regulatory mechanisms or capture effects in target organs such as lung epithelium (37). Constraints inherent to working with PBMC may have contributed to the 35 GWAS lead SNPs that did not colocalize in our analysis. Sample sizes differed between cohorts (TG1-3, n = 737; TG4-5, n = 294), and longitudinal attrition further reduced the TG4-5 cohort at later timepoints, which may limit detection of late-stage regulatory effects in patients with the most critical disease due to insufficient statistical power. IMPACC participants were largely unvaccinated, so the cis-eQTL regulatory effects reported here may have limited generalizability beyond this early-pandemic, unvaccinated, hospitalized cohort. Despite the biological and clinical plausibility of the genetic mechanisms revealed by our Bayesian colocalization analyses, functional studies are required to confirm the statistical findings linking these GWAS-identified SNPs to the candidate causal genes.

In summary, cis-regulatory effects of critical COVID-19 GWAS lead SNPs vary across clinical severity and disease phase, and many are detectable only when cis-eQTL study design incorporates severity stratification and longitudinal sampling. Examples include severity-cohort-specific regulation at *TMIGD2*, where the protective allele upregulated expression exclusively in patients with prolonged-critical/fatal illness (TG4-5) during the early acute phase. Candidate causal genes identified through this approach generate testable hypotheses about disease mechanisms and potential therapeutic targets for critical COVID-19.

## Materials and Methods

### Ethics

Twelve institutions elected to conduct the study under the public health surveillance exception [45CFR46.102(l)(2)], while three sites with prior IRB-approved biobanking protocols elected to integrate and conduct IMPACC under their institutional protocols (University of Texas at Austin, IRB 2020-04-0117; University of California San Francisco, IRB 20-30497; Case Western reserve university, IRB STUDY20200573) with informed consent requirements. Participants enrolled under the public health surveillance exception were provided information sheets describing the study, samples to be collected, and plans for data deidentification and usage; individuals declining participation were not enrolled. Participants received no compensation while hospitalized but were offered compensation for outpatient follow-up (38).

### IMPACC Study Design and Participants

IMPACC (21, 38) is a prospective, multicenter longitudinal cohort study of hospitalized adults with laboratory-confirmed COVID-19 (https://clinicaltrials.gov/study/NCT04378777). Between May 5, 2020 and March 19, 2021, a total of 1,164 participants were enrolled across 15 research centers encompassing 20 hospital sites throughout the United States. Participants were recruited within 48 hours of hospital admission. Inclusion criteria were as follows: (1) age ≥18 years; (2) laboratory-confirmed SARS-CoV-2 infection by RT-PCR or antigen test; and (3) hospitalization for COVID-19 illness with symptoms (21). Exclusion criteria included pregnancy. This cohort demonstrated substantial demographic and clinical diversity; detailed participant characteristics are presented in prior reports (21, 38). Concurrent enrollment in other observational or interventional protocols was permitted. IMPACC represents unvaccinated individuals as enrollment was completed prior to COVID-19 vaccine availability in the United States. Longitudinal clinical assessments and biospecimen collection occurred at up to 10 timepoints: enrollment (V1; within 48 hours of admission), acute phase (V2-6, at days: 4, 7, 14, 21, and 28), and convalescent phase (V7-10, at months: 3, 6, 9, and 12 post-discharge). This current analysis focuses on the subset of 921 participants (79.1% of the total cohort) passing genotype quality control and with PBMC RNA-sequencing data available at visit 1.

### Clinical trajectory classification

Clinical trajectory groups (TGs 1-5) were defined within IMPACC by latent class modeling of longitudinal WHO ordinal scale assessments collected during the first 28 days post-admission, yielding five trajectories from rapid recovery (TG1, n = 204) to death (TG5, n = 99) (21). Following prior IMPACC analyses (22, 23), we aggregated these into two cohorts for genetic analysis: a rapid-recovery cohort (TG1-3, n = 737), comprising patients who recovered within 28 days of hospital admission, and a prolonged-critical/fatal cohort (TG4-5, n = 294), comprising patients with prolonged hospitalization or death within 28 days. This binary stratification is clinically meaningful and provides sufficient sample sizes per stratum for cis-eQTL mapping. All IMPACC participants across all five trajectory groups correspond to the critical illness phenotype defined in the GenOMICC GWAS (11), which serves as the disease-trait reference summary statistics for our colocalization analyses.

### Genetic Data Generation and Quality Control

Genomic DNA was extracted from whole blood and genotyped using the Illumina Global Diversity Array at the Yale Center for Genomic Analysis. Quality control was performed using PLINK v1.9 (39), retaining samples and variants with >95% call rates, MAF >1%, and Hardy-Weinberg equilibrium p>10⁻⁶. We excluded related individuals (pi-hat > 0.1875), samples with discordance between clinical and genetically inferred sex, and population outliers identified through ADMIXTURE analysis (40), resulting in a genotype dataset of 1,031 participants. Genotypes were phased using ShapeIt4 (41) and imputed with Beagle5.4 (42), retaining variants with imputation quality R² ≥0.7 (21,748,405 variants). To address population stratification, we aligned our samples with the 1000 Genomes Project (Phase 3) and calculated ancestry-specific allele frequencies. We performed ancestry-stratified filtering, removing variants with MAF < 5% within each ADMIXTURE-defined stratum to minimize false positives driven by population substructure. Robust principal components calculated via PC-AiR (in GENESIS package v2.36) are included as covariates in all genetic models.

### Statistical Analysis

#### Transcriptomic Data Processing and Longitudinal Design

Gene expression was normalized following the GTEx Consortium protocol (https://gtexportal.org/home/methods). Briefly, read counts were TMM-normalized (edgeR v4.4.2) (43), filtered to retain genes with expression ≥0.1 TPM and ≥6 reads in ≥20% of samples, and inverse normal transformed. Non-coding genes were removed. To capture context-dependent regulation, we analyzed each visit-trajectory stratum independently (excluding Visit 5 for TG1-3 due to low sample size, n = 10). PCA-based outliers (>4 SD in PC1-2) were removed per stratum. Sample sizes ranged from n = 660 (Visit 1 for TG1-3) to n = 41 (Visit 10 for TG4-5) due to discharge and mortality.

#### Cis-eQTL Mapping and Multiple Testing Correction

All analyses were performed in R v4.3.3. Linear regression was performed using MatrixEQTL v2.3 with an additive genetic model (44) testing all variants within ±1 Mb of 58 established risk loci (299,380 variants). To control for confounding heterogeneity, models included age at hospital admission, sex, population substructure (genotype PCs via PC-AiR, GENESIS package v2.36, with the number of retained components determined by nScree from the nFactors package v2.4.1.1) (45, 46), and technical gene expression variation (PCs selected via Buja-Eyuboglu permutation test implemented via runBE from the PCAForQTL package v0.1.0) (47). The number of latent factors was optimized algorithmically for each severity-by-visit stratum (typically 23-49 covariates) to maximize variance explained without overfitting. Multiple-testing correction was performed using eigenMT (https://github.com/joed3/eigenMT) (24), which adjusts each gene’s association P-value for the effective number of independent variants in its cis-window. This accounts for linkage disequilibrium among variants and closely approximates permutation-based correction. We applied an eigenMT-adjusted P < 0.01 threshold for each tested gene within each stratum, treating each visit-trajectory combination as an independent experiment.

#### Bayesian Colocalization

We performed colocalization between IMPACC cis-eQTLs and GenOMICC GWAS summary statistics (Release 3: 23,707 cases and 1,075,129 controls) (11) using coloc.abf from the coloc package v5.2.3 (25). Summary statistics were harmonized to align effect alleles, and ambiguous palindromic SNPs (MAF 0.4-0.6) were excluded to ensure directional consistency of effect sizes. For every statistically significant IMPACC cis-eQTL at each visit-trajectory stratum (eigenMT-adjusted P < 0.01), we tested colocalization within a ±250 kb window. We utilized a conservative prior probability of a shared causal variant (P12 of 5.5×10⁻⁶) and defined colocalization as a posterior probability PP.H4 > 0.5 (with independent signal probability PP.H3 < 0.5), consistent with thresholds used by GenOMICC (10, 11).

#### Statistical Comparison of Candidate-Gene Detection Between Analyses

To test whether the severity-stratified, longitudinal analysis identified candidate causal genes at more of the 58 tested lead SNPs than the unstratified hospital admission analysis, each lead SNP was classified as colocalized (PP.H4 > 0.5) or not colocalized under each analysis. Then, the two analyses were compared as paired binary outcomes using McNemar’s test. The exact test was applied, computed as a two-sided exact binomial test on the discordant pairs (binom.test, R v4.3.3), with two-sided p < 0.05 considered significant.

## Data Availability

Data used for this study are available through the ImmPort database (accession SDY1760) upon approval of data access. The IMPACC genotypes are available through dbGaP (accession phs002686.v3.p2). All analysis code is available at: https://bitbucket.org/kleinstein/impacc-public-code/src/master/eqtl_genetics_manuscript/

## Acknowledgments

We thank the participants of the study for their voluntary enrollment and contribution of samples for this work. See the supplement for details on the IMPACC Network. We thank the Yale Center for Genomic Analysis (YCGA) which greatly facilitated this work. We acknowledge the assistance of the following individuals: Sanya Thomas, Mitchell Cooney, Shun Rao, Sofia Vignolo, and Elena Morrocchi (all from the CDCC); Arash Naeim, Marianne Bernardo, Sarahmay Sanchez, Shannon Intluxay, Clara Magyar, Jenny Brook, Estefania Ramires-Sanchez, Megan Llamas, Claudia Perdomo, Clara E. Magyar, Jennifer A. Fulcher, Clara Frydman, and Brunilda Balliu (all from the David Geffen School of Medicine at UCLA); members of the UCLA Center for Pathology Research Services and the Pathology Research Portal; Dimitri Duvilaire, Maxine Kuang, Denise Shepherd, and Chris Cotsapas (all from the Yale School of Medicine). We thank the leadership of Boston Children’s Hospital including Drs. Wendy Chung, Gary Fleisher, Nancy Andrews, and Kevin Churchwell for their support for the Precision Vaccines Program. Dr. Augustine’s and Becker’s co-authorship of this report does not necessarily represent the official views of the National Institute of Allergy and Infectious Diseases, the National Institutes of Health or any other agency of the United States Government.

## Competing Interests

### IMPACC Network

The Icahn School of Medicine at Mount Sinai has filed patent applications related to SARS-CoV-2 serological assays, NDV-based SARS-CoV-2 vaccines, influenza virus vaccines, and therapeutics, listing Florian Krammer and Viviana Simon as co-inventors. Mount Sinai has spun out Kantaro to market SARS-CoV-2 serological tests and Castlevax to develop SARS-CoV-2 vaccines, with Florian Krammer as co-founder and scientific advisory board member of Castlevax. Florian Krammer has consulted for Merck, Curevac, Seqirus, GSK, Pfizer, 3rd Rock Ventures, Sanofi, Gritstone, and Avimex. His laboratory collaborates with Dynavax on influenza vaccine development and with VIR on influenza virus therapeutics development. Ofer Levy is a named inventor on patents held by Boston Children’s Hospital related to vaccine adjuvants and human in vitro platforms that model vaccine action. His laboratory has received research support from GlaxoSmithKline (GSK) and Pfizer, he is a consultant to GSK and Sanofi, and a co-founder and advisor to *ARMR Sciences*, which develops immunotherapeutics to prevent opioid overdose. Charles Cairns consults for bioMérieux and receives grant funding from the Bill & Melinda Gates Foundation. James A. Overton is a consultant at Knocean Inc. Jessica Lasky-Su is a scientific advisor for Precion Inc. Scott R. Hutton, Greg Michelloti, and Kari Wong are employees of Metabolon Inc. Vicki Seyfer-Margolis is employed by MyOwnMed. Nadine Rouphael reports grants or contracts with Merck, Sanofi, Pfizer, Vaccine Company, and Immorna. She has served on data safety monitoring and advisory boards for Moderna, AZ, Sanofi, Seqirus, Pfizer, EMMES, ICON, BARDA, Imunon, and CyanVac. She has received travel support from Sanofi and Moderna and honoraria from Virology Education and Krog Consulting. Chris Cotsapas is employed by Vesalius Therapeutics. Adeeb Rahman is employed by Immunai Inc. Steven Kleinstein consults for Peraton related to the ImmPort data repository. Nathan Grubaugh consults for Tempus Labs and the National Basketball Association. Akiko Iwasaki consults for 4BIO, Blue Willow Biologics, Revelar Biotherapeutics, RIGImmune, Xanadu Bio, and Paratus Sciences. Monica Kraft receives research funding from NIH, ALA, Sanofi, and AstraZeneca for asthma research. She consults for AstraZeneca, Sanofi, Chiesi, and GSK for severe asthma and is co-founder and CMO of RaeSedo, Inc., developing peptidomimetics for inflammatory lung disease. Esther Melamed receives research funding from Babson Diagnostics, honoraria from the Multiple Sclerosis Association of America, and has served on advisory boards for Genentech, Horizon, Teva, and Viela Bio. Carolyn Calfee receives research funding from NIH, FDA, DOD, Roche-Genentech, and Quantum Leap Healthcare Collaborative and consults for Janssen, Vasomune, Gen1e Life Sciences, NGMBio, and Cellenkos. Wade Schulz has collaborated with the Shenzhen Center for Health Information and the National Center for Cardiovascular Diseases in Beijing, is a technical consultant for Hugo Health, co-founder of Refactor Health, and has received grants from Merck and Regeneron Pharmaceuticals for COVID-19 research. Grace A. McComsey receives research grants from Redhill, Cognivue, Pfizer, and Genentech and consults for Gilead, Merck, and ViiV/GSK. Linda N. Geng receives research funding from Pfizer, Inc., through her institution. Catherine Hough receives research support from NIH and CDC. David Hafler has received research funding from Bristol-Myers Squibb, Novartis, Sanofi, and Genentech and consults for Bayer Pharmaceuticals, Repertoire Inc., Bristol Myers Squibb, Compass Therapeutics, EMD Serono, Genentech, Novartis Pharmaceuticals, and Sanofi Genzyme.

## The IMPACC Network

**National Institute of Allergy and Infectious Diseases, National Institute of Health, Bethesda, MD 20814, USA**

Patrice M. Becker, Alison D. Augustine, Steven M. Holland, Lindsey B. Rosen, Serena Lee, Tatyana Vaysman

**Clinical and Data Coordinating Center (CDCC), Precision Vaccines Program, Boston Children’s Hospital, Harvard Medical School, Boston, MA 02115, USA**

Al Ozonoff, Joann Diray-Arce, Jing Chen, Alvin T. Kho, Carly E. Milliren, Annmarie Hoch, Ana C. Chang, Kerry McEnaney, Caitlin Syphurs, Brenda Barton, Claudia Lentucci, Maimouna D. Murphy, Mehmet Saluvan, Tanzia Shaheen, Shanshan Liu, Marisa Albert, Arash Nemati Hayati, Robert Bryant, James Abraham, Mitchell Cooney, Meagan Karoly

**Benaroya Research Institute, University of Washington, Seattle, WA 98101, USA**

Matthew C. Altman, Naresh Doni Jayavelu, Scott Presnell, Bernard Kohr, Tomasz Jancsyk, Azlann Arnett

**La Jolla Institute for Immunology, La Jolla, CA 92037, USA**

Bjoern Peters, James A. Overton, Randi Vita, Kerstin Westendorf

**Knocean Inc. Toronto, ON M6P 2T3, Canada**

James A. Overton

**Precision Vaccines Program, Boston Children’s Hospital, Harvard Medical School, Boston, MA 02115, USA**

Ofer Levy, Hanno Steen, Patrick van Zalm, Benoit Fatou, Kinga K. Smolen, Arthur Viode, Simon van Haren, Meenakshi Jha, David Stevenson, Athena N. Nguyen, Alec L. Plotkin, Sanya Thomas, Boryana Petrova, Naama Kanarek

**Brigham and Women’s Hospital, Harvard Medical School, Boston, MA 02115, USA**

Lindsey R. Baden, Kevin Mendez, Jessica Lasky-Su, Alexandra Tong, Rebecca Rooks, Michael Desjardins, Amy C. Sherman, Stephen R. Walsh, Xhoi Mitre, Jessica Cauley, Xiaofang Li, Bethany Evans, Christina Montesano, Jose Humberto Licona, Jonathan Krauss, Nicholas C. Issa, Jun Bai Park Chang, Natalie Izaguirre

**Metabolon Inc, Morrisville, NC 27560, USA**

Scott R. Hutton, Greg Michelotti, Kari Wong

**Prevention of Organ Failure (PROOF) Centre of Excellence, University of British Columbia, Vancouver, BC V6T 1Z3, Canada:**

Scott J. Tebbutt, Casey P. Shannon

**Case Western Reserve University and University Hospitals of Cleveland, Cleveland, OH 44106, USA:**

Rafick-Pierre Sekaly, Slim Fourati, Grace A. McComsey, Paul Harris, Scott Sieg, George Yendewa, Mary Consolo, Heather Tribout, Susan Pereira Ribeiro

**Drexel University, Tower Health Hospital, Philadelphia, PA 19104, USA**

Charles B. Cairns, Elias K. Haddad, Michele A. Kutzler, Mariana Bernui, Gina Cusimano, Jennifer Connors, Kyra Woloszczuk, David Joyner, Carolyn Edwards, Edward Lee, Edward Lin, Nataliya Melnyk, Debra L. Powell, James N. Kim, I. Michael Goonewardene, Brent Simmons, Cecilia M. Smith, Mark Martens, Brett Croen, Nicholas C. Semenza, Mathew R. Bell, Sara Furukawa, Renee McLin, George P. Tegos, Brandon Rogowski, Nathan Mege, Kristen Ulring, Pam Schearer, Judie Sheidy, Crystal Nagle

**MyOwnMed Inc., Bethesda, MD 20817, USA**

Vicki Seyfert-Margolis

**Emory School of Medicine, Atlanta, GA 30322, USA**

Nadine Rouphael, Steven E. Bosinger, Arun K. Boddapati, Greg K. Tharp, Kathryn L. Pellegrini, Brandi Johnson, Bernadine Panganiban, Christopher Huerta, Evan J. Anderson, Hady Samaha, Jonathan E. Sevransky, Laurel Bristow, Elizabeth Beagle, David Cowan, Sydney Hamilton, Thomas Hodder, Amer Bechnak, Andrew Cheng, Aneesh Mehta, Caroline R. Ciric, Christine Spainhour, Erin Carter, Erin M. Scherer, Jacob Usher, Kieffer Hellmeister, Laila Hussaini, Lauren Hewitt, Nina Mcnair, Susan Pereira Ribeiro, Sonia Wimalasena

**Icahn School of Medicine at Mount Sinai, New York, NY 10029, USA**

Ana Fernandez-Sesma, Viviana Simon, Florian Krammer, Harm van Bakel, Seunghee Kim-Schulze, Ana Silvia Gonzalez-Reiche, Jingjing Qi, Brian Lee, Juan Manuel Carreño, Gagandeep Singh, Ariel Raskin, Johnstone Tcheou, Zain Khalil, Adriana van de Guchte, Keith Farrugia, Zenab Khan, Geoffrey Kelly, Komal Srivastava, Lily Q. Eaker, Maria C. Bermúdez-González, Lubbertus C.F. Mulder, Katherine F. Beach, Miti Saksena, Deena Altman, Erna Kojic, Levy A. Sominsky, Arman Azad, Dominika Bielak, Hisaaki Kawabata, Temima Yellin, Miriam Fried, Leeba Sullivan, Sara Morris, Giulio Kleiner, Daniel Stadlbauer, Jayeeta Dutta, Hui Xie, Manishkumar Patel, Kai Nie, Brian Monahan

**Immunai Inc., New York, NY 10016, USA**

Adeeb Rahman

**Oregon Health & Science University, Portland, OR 97239, USA**

William B. Messer, Catherine L. Hough, Sarah A.R. Siegel, Peter E. Sullivan, Zhengchun Lu, Amanda E. Brunton, Matthew Strand, Zoe L. Lyski, Felicity J. Coulter, Courtney Micheletti

**Stanford University School of Medicine, Palo Alto, CA 94305, USA**

Holden Maecker, Bali Pulendran, Kari C. Nadeau, Yael Rosenberg-Hasson, Michael Leipold, Natalia Sigal, Angela Rogers, Andrea Fernandes, Monali Manohar, Evan Do, Iris Chang, Alexandra S. Lee, Catherine Blish, Henna Naz Din, Jonasel Roque, Linda N. Geng, Maja Artandi, Mark M. Davis, Neera Ahuja, Samuel S. Yang, Sharon Chinthrajah, Thomas Hagan, Tyson H. Holmes, Koji Abe

**David Geffen School of Medicine at the University of California Los Angeles, Los Angeles CA 90095, USA**

Elaine F. Reed, Joanna Schaenman, Ramin Salehi-Rad, Adreanne M. Rivera, Harry C. Pickering, Subha Sen, David Elashoff, Dawn C. Ward, Jenny Brook, Estefania Ramires-Sanchez, Megan Llamas, Claudia Perdomo, Clara E. Magyar, Jennifer Fulcher

**University of California San Francisco, San Francisco, CA 94115, USA**

David J. Erle, Carolyn S. Calfee, Carolyn M. Hendrickson, Kirsten N. Kangelaris, Viet Nguyen, Deanna Lee, Suzanna Chak, Rajani Ghale, Ana Gonzalez, Alejandra Jauregui, Carolyn Leroux, Luz Torres Altamirano, Ahmad Sadeed Rashid, Andrew Willmore, Prescott G. Woodruff, Matthew F. Krummel, Sidney Carrillo, Alyssa Ward, Charles R. Langelier, Ravi Patel, Michael Wilson, Ravi Dandekar, Bonny Alvarenga, Jayant Rajan, Walter Eckalbar, Andrew W. Schroeder, Gabriela K. Fragiadakis, Alexandra Tsitsiklis, Eran Mick, Yanedth Sanchez Guerrero, Christina Love, Lenka Maliskova, Michael Adkisson, Aleksandra Leligdowicz, Alexander Beagle, Arjun Rao, Austin Sigman, Bushra Samad, Cindy Curiel, Cole Shaw, Gayelan Tietje-Ulrich, Jeff Milush, Jonathan Singer, Joshua J. Vasquez, Kevin Tang, Legna Betancourt, Lekshmi Santhosh, Logan Pierce, Maria Tecero Paz, Michael Matthay, Neeta Thakur, Nicklaus Rodriguez, Nicole Sutter, Norman Jones, Pratik Sinha, Priya Prasad, Raphael Lota, Saurabh Asthana, Sharvari Bhide, Tasha Lea, Yumiko Abe-Jones

**Yale School of Medicine, New Haven, CT 06510, USA**

David A. Hafler, Ruth R. Montgomery, Albert C. Shaw, Steven H. Kleinstein, Jeremy P. Gygi, Dylan Duchen, Shrikant Pawar, Anna Konstorum, Ernie Chen, Chris Cotsapas, Xiaomei Wang, Charles Dela Cruz, Akiko Iwasaki, Subhasis Mohanty, Allison Nelson, Yujiao Zhao, Shelli Farhadian, Hiromitsu Asashima, Omkar Chaudhary, Andreas Coppi, John Fournier, M. Catherine Muenker, Khadir Raddassi, Michael Rainone, William Ruff, Syim Salahuddin, Wade L. Shulz, Pavithra Vijayakumar, Haowei Wang, Esio Wunder Jr., H. Patrick Young, Albert I. Ko, Gisela Gabernet

**Yale School of Public Health, New Haven, CT 06510, USA**

Denise Esserman, Leying Guan, Anderson Brito, Jessica Rothman, Nathan D. Grubaugh, Kexin Wang, Leqi Xu

**Baylor College of Medicine and the Center for Translational Research on Inflammatory Diseases, Houston, TX 77030, USA**

David B. Corry, Farrah Kheradmand, Li-Zhen Song, Ebony Nelson

**Oklahoma University Health Sciences Center, Oklahoma City, OK 73104, USA**

Jordan P. Metcalf, Nelson I. Agudelo Higuita, Lauren A. Sinko, J. Leland Booth, Douglas A. Drevets, Brent R. Brown

**University of Arizona, Tucson AZ 85721, USA**

Monica Kraft, Christian Bime, Jarrod Mosier, Heidi Erickson, Ron Schunk, Hiroki Kimura, Michelle Conway, Dave Francisco, Allyson Molzahn, Connie Cathleen Wilson, Ron Schunk, Trina Hughes, Bianca Sierra

**University of Florida, Gainesville, FL 32611, USA**

Mark A. Atkinson, Scott C. Brakenridge, Ricardo F. Ungaro, Brittany Roth Manning, Lyle Moldawer

**University of Florida, Jacksonville, FL 32218, USA**

Jordan Oberhaus, Faheem W. Guirgis

**University of South Florida, Tampa FL 33620, USA**

Brittney Borresen, Matthew L. Anderson

**The University of Texas at Austin, Austin, TX 78712, USA**

Lauren I. R. Ehrlich, Esther Melamed, Cole Maguire, Dennis Wylie, Justin F. Rousseau, Kerin C. Hurley, Janelle N. Geltman, Nadia Siles, Jacob E. Rogers, Pablo Guaman Tipan

---

**Fig. S1.**
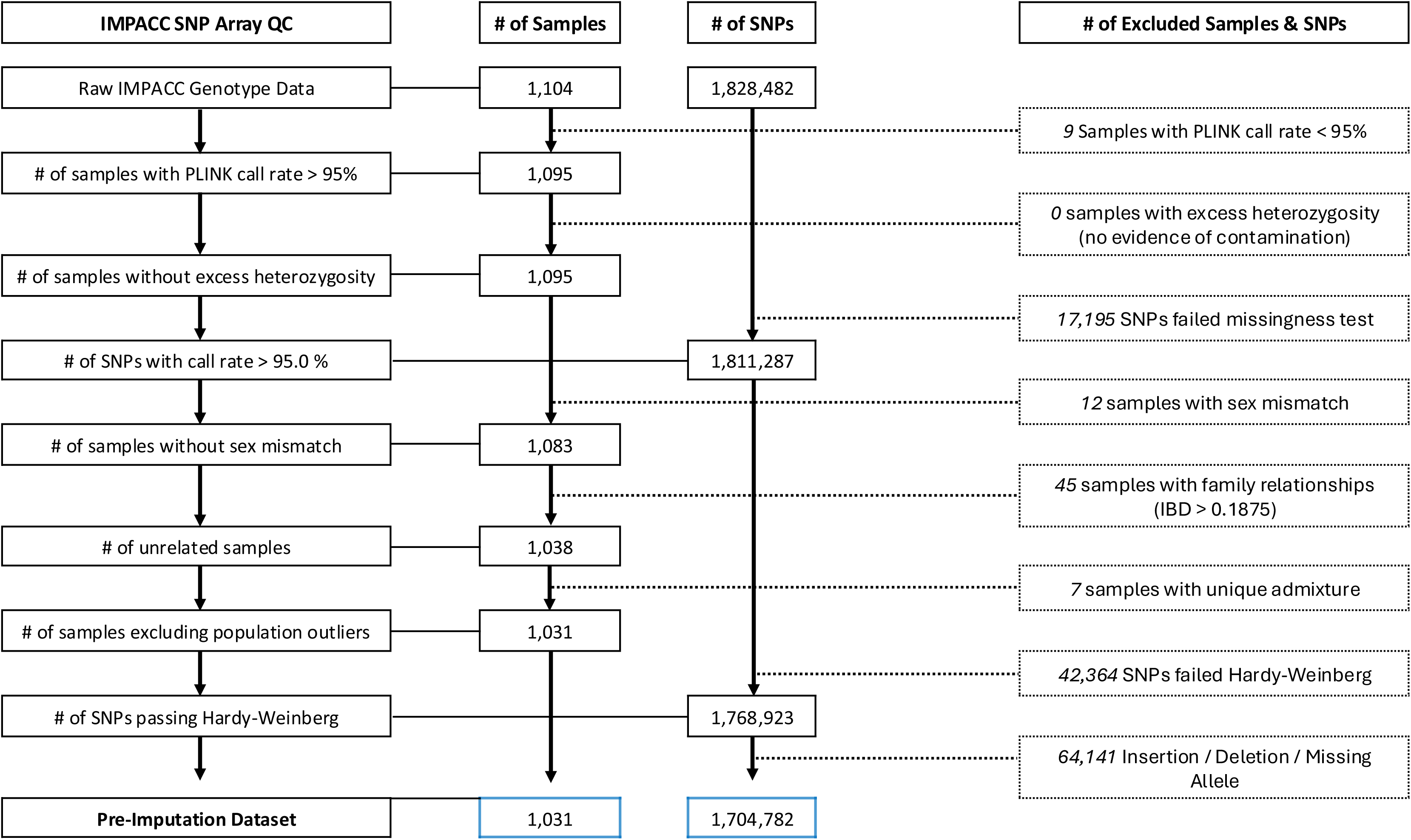
Sample and variant quality-control workflow for the IMPACC genotype data. Flow diagram of the quality-control (QC) pipeline applied to Illumina Global Diversity Array genotypes, showing retained samples (column 2) and SNPs (column 3) at each step from the raw call set to the pre-imputation dataset, with the corresponding reason for exclusions at right. Samples are excluded for a call rate < 95%, excess heterozygosity indicative of contamination, discordance between clinical and genetically inferred sex, cryptic relatedness (IBD > 0.1875), or population-outlier status by ADMIXTURE. SNPs are excluded for a call rate < 95%, departure from Hardy-Weinberg equilibrium (P < 1 × 10^−6^), or insertion, deletion, or missing-allele status. Of the initial 1,104 participants with genotype data and 1,828,482 SNPs, 1,031 participants and 1,704,782 SNPs pass QC to form the pre-imputation dataset.

**Fig. S2.**
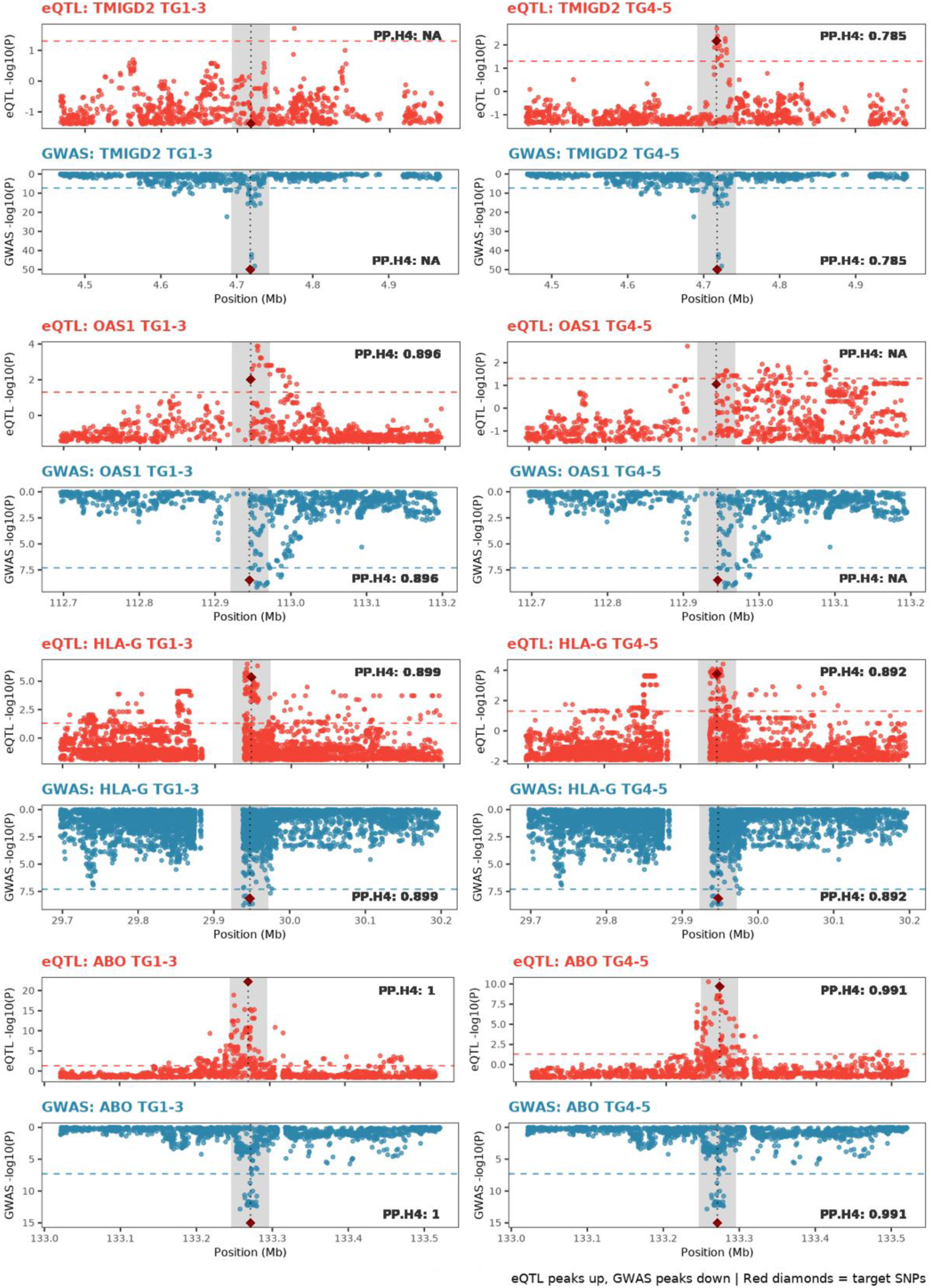
Regional association (mirror) plots of cis-eQTL and GWAS signals at the four representative loci from Fig. 4. For each locus (*TMIGD2, OAS1, HLA-G, ABO*), paired plots span a 500-kb window (±250 kb around the lead SNP) in the rapid-recovery (TG1-3, left) and prolonged-critical/fatal (TG4-5, right) cohorts. Within each pair, the IMPACC cis-eQTL association (top) is plotted above the GenoMICC critical COVID-19 GWAS association (bottom, inverted). The y-axis shows -log10(P) and the x-axis genomic position (Mb, GRCh38). Point color denotes the association (red = cis-eQTL; blue = GWAS). The dark red diamond marks the reported GWAS lead SNP, the dotted vertical line its position, and grey shading a ±25-kb (50-kb) window centered on the lead SNP. Dashed horizontal lines mark reference levels for significance (red, cis-eQTL eigenMT-adjusted P = 0.05; blue, GWAS genome-wide significance, P = 5 × 10⁻⁸). The posterior probability of a shared causal variant (PP.H4) from Bayesian colocalization is annotated in each panel and reported as NA where no colocalization with PP.H4 > 0.5 was detected. The cis-eQTL panels correspond to Visit 1 (*TMIGD2* and *HLA-G* in both cohorts, and *OAS1* in TG4-5), Visit 6 (*OAS1* in TG1-3), and Visit 3 (ABO in both cohorts), in each case the visit of maximal PP.H4 for that locus and cohort, or Visit 1 where none was detected. Colocalization is cohort-specific for *TMIGD2* (TG4-5 only, PP.H4 = 0.785) and *OAS1* (TG1-3 only, PP.H4 = 0.896), present in both cohorts for *HLA-G* (PP.H4 = 0.899 and 0.892), and constitutive in both for *ABO* (PP.H4 = 1.00 and 0.991).

